# Predicting the Dengue Epidemic in Guangzhou, China by Meteorological Parameter Methods

**DOI:** 10.1101/2020.04.23.20076448

**Authors:** Jing Chen, Kang-Kang Liu, Hui Xiao, Gang Hu, Xiang Xiao, Yu Lin, Qian Yue, Yan Han, Xiao-Fei Mao, Guang-Hui Dong, Jin Bu

**Affiliations:** Institute of Tropical and Marine Meteorology, China Meteorological Administration, Guangzhou, Chuangdong 510080, China; School of Atmospheric Science, Sun Yat-sen University, Guangzhou, Guangdong 519082, China; Guangzhou Key Laboratory of Environmental Pollution and Health Risk Assessment, Department of Occupational and Environmental Health, School of Public Health, Sun Yat-sen University, Guangzhou, Guangdong 510080, China; School of Agriculture, Sun Yat-sen University, Guangzhou, Guangdong 510275, China; Guangzhou South China Biomedical Research Institute co., Ltd, Guangzhou, Guangdong 510275, China; Hospital for Skin Diseases (Institute of Dermatology), Chinese Academy of Medical Sciences and Peking Union Medical College, Nanjing, Jiangsu 210042, China

**Author notes:** Corresponding authors: Jin Bu,; Guang-Hui Dong,; Xiao-Fei Mao,.

**Keywords:** Dengue, Epidemic, Meteorological Parameter, Probabilistic Forecast, Sliding Accumulated Temperature, Imported Case, Sporadic Case, Concentrated patients, Temporal Characteristics, Spatial Distribution

## Abstract

This study was aimed to determine dengue season, and further establish a prediction model by meteorological methods. The dengue and meteorological data were collected from Guangdong Meteorological Bureau and Guangdong Provincial Center for Disease Prevention and Control, respectively. We created a sliding accumulated temperature method to accurately calculate the beginning and ending day of dengue season. Probabilistic Forecast model was derived under comprehensive consideration of various weather processes including typhoon, rainstorm, and so on. We found: 1) The dengue fever season enters when effective accumulated temperature of a continuing 45 days (T_45_) ≥0 °C, and it finishes when effective accumulated temperature of a continuing 6 days (T_6_) <0 °C. 2) A Probabilistic Forecast Model for dengue epidemic was established with good forecast effects, which were verified by the actual incidence of dengue in Guangzhou. The Probabilistic Forecast Model provides markedly improved forecasting techniques for dengue prediction.

## Introduction

Dengue has become an increasing public health threat around the world due to the highest incidence and the fastest spread rate, serious health consequences including death, lack of effective treatment and vaccine (*1*). In the past 15 years, the global incidence of dengue fever has increased by 70%, involving in 128 countries and regions. There are about 390 million people annually suffering from dengue fever, of which ~96 million are symptomatic(*2*).

In Chinese mainland, dengue cases have been reported every year since 1997, and become one of the critical infectious diseases to be prevented and controlled. Guangzhou city is the hardest hit area of dengue with 90% cases reported of the whole country. It may be related to the geographical and climatic environment of Guangzhou, which is located in south China and closed to the Southeast Asia countries, and *Aedes albopictus* and *Aedes aegypti* are the main vectors in Guangzhou(*3*). In the history of Guangzhou, there were 3 times with more than 1,000 cases reported one year (2006, 2013, and 2014). One outbreak happened in 2014, and affected 20 cities over 42,335 cases reported(*3, 4*).

**Supplementary Figure 1** shows the Guangdong-Hong Kong-Macao Greater Bay Area, and the geographic distribution of dengue cases in 2014, which presented as orange red area with a large number of cases concentrated. Guangzhou is the center of the Guangdong-Hong Kong-Macao greater bay area, meaning that the six core cities in the greater bay area, namely Hong Kong, Shenzhen, Foshan, Zhuhai, and Macao, are facing the threat of dengue transmission. With the deepening of regional cooperation in the urban agglomeration of the greater bay area and the acceleration of population flow, the greater bay area will be more favorable for the spread of dengue. Dengue prevention and control is in a greater difficulty. The government annually spends nearly 200 million yuan on disease control, but the results were far from satisfying. The whole urban agglomeration of the greater bay area and surrounding areas is facing more severe challenges than in 2014. Therefore, it is urgent to establish an early warning and predicting system with high-precision for dengue.

Climate conditions have been identified as important factors affecting the transmission of dengue(*5-7*), for example, temperature affacts the biting activity and the distribution of the *Aedes aegypti*(*8*) and virus development(*9*). Humidity also has an impact on the population density of female mosquitoes. Furthermore, Mosquitoes live longer and disperse further under high relative humidity conditions(*1*), and distinct seasonal pattern in the outbreaks of dengue viruses around the world(*1*) were consistent with the beginning and end of rainy season. An increase in dengue was proved in association with an increase in average monthly rainfall and average monthly maximum temperature(*7, 10*).

As meteorological factors play key roles in the epidemiology of dengue, we aimed to develop tailored climate-based forecasting models of dengue fever for Guangzhou city. This study took the collaboration between meteorology and public health one step further, by co-developing a dengue early probabilistic forecast model using meteorological parameters, provides markedly improved forecasting techniques for disease prediction, and could potentially be operationalised as a meteorological service for the public health sector.

## Materials and Methods

### 1. Meteorological data

We collected the meteorological data from January 1, 2005 and December 31, 2016 from Guangdong Meteorological Bureau, including daily average temperature, daily precipitation, daily average humidity and daily weather phenomena (continuous precipitation days and continuous no precipitation days).

### 2. Dengue case data

We collected the dengue data between January 1, 2005 and December 31, 2016 from Guangdong Provincial Center for Disease Prevention and Control, including the number of cases reported per day with the onset time and the detailed address of the patients, and whether the case was imported.

Imported cases are defined as those who have traveled to dengue endemic regions and been bitten by mosquitoes less than 15 days before the symptom onset(*11*).

### 3. Analysis of imported dengue cases on dengue epidemic

According to the onset time information and residential address of the imported cases, a continuous time series was formed from January 1, 2005 to December 31, 2016. And then, high-precision geographic grid points were used to conduct spatiotemporal correlation analysis of the imported cases by the following steps:

1. Time correlation analysis: If the onset time of case was before entry of Guangzhou, the entry time was taken as the starting point for the analysis; If the onset time was after entry, the onset time was taken as the starting point. The density of dengue patients was calculated daily from starting point.
2. Spatial correlation analysis: the residential address of the imported case was taken as the center of the circle, and the density of patients in different radii was calculated every 100 meters within the range from 0 to 20 km using 100 meters apart. The distance of non-imported case from the imported case was calculated according to **Formula 1**.
3. Spatiotemporal correlation analysis: the spatiotemporal influence of the imported cases on around cases was determined both by step 1 and step 2. According to the flight distance of mosquitoes, it is generally believed that the range of the mosquito’s activity is mainly within 100 m of its birthplace, and the maximum is no more than 1 km(*12*). We calculated the potential incidence of dengue in this high risk area.

#### Formula 1

The distance calculation formula from D_*i*_ to D_*i+1*_:

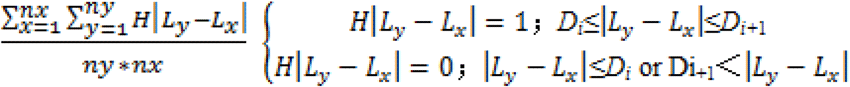

Note: *x* is the imported case, *y* is the non-imported case, *L*_*x*_ is the location of the imported case, *L*_*y*_ and is the location of the non-imported case.

### 4. The interpolation of climatic data of dengue cases

The longitude and latitude of automatic stations in Guangzhou City and the residential address of dengue cases were collected from Guangzhou Meteorological Bureau. We interpolated the meteorological data of the location of each dengue case at the date of diagnosis by **Formula 2**, and the calculated meteorological data were used for the further analyses of relationship between meteorology and dengue, and to establish the probabilistic forecast equation.

#### Formula 2

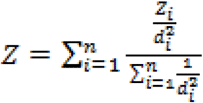

Note: *n* is the sample number, *Z*_*i*_ is the climatic data of sampling site, and *d*_*i*_ is the distance from *i* to the interpolated site. When the sampling site and the interpolated site overlap with, the sampling site weight is 1, and the Z value of interpolated site is equal to the sampling site.

### 5. The sliding accumulated temperature method

In Guangzhou, when the temperature reaches 17 °C, mosquitoes begin to move(*13*), so we set 17 °C as the base temperature with every 0.5 °C as the interval, and effective temperature (T) can be calculated by **Formula 3**:

#### Formula 3

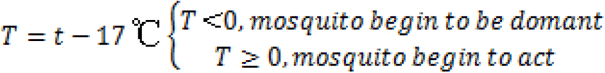

Note: *t* is the mean temperature of current day.

We defined cumulative effective temperature of a continuing *n* days as sliding accumulated temperature (T_*n*_), when T_*n*_ ≥0 °C, mosquitoes can complete incubation, and their concentration is conducive to the occurrence and transmission of dengue virus. When the effective accumulated temperature was less than 0 for *n’* consecutive days, which is defined as T_*n’*_, the hatching of mosquitoes began to be significantly inhibited, and the mosquito activity sharply decreased. Mosquitoes disappearing from the nature in large numbers means dengue fever entered a low incidence period.

### 6. Quantification of weather phenomena/process by meteorological indexes

Major weather phenomena/process in Guangzhou include typhoons, strong precipitation, high temperature, cold air mass, drought, typhoon, etc. When one weather process continues for a period of time, the mosquito will be obviously affected by the weather and the growth environment, but how to accurately evaluate the impact of weather process on mosquito? We firstly screened meteorological data to identify all of the weather phenomena and determined their influence on mosquito survival, reproduction, and activity. And then we used the following common meteorological indexes to quantify weather phenomena/process.

1. Temperature (unit: °C, symbol T) is used to indicate the effects of high temperature and low temperature;
2. Relative humidity (unit: %, symbol τ, is available under various weather phenomena;
3. Daily precipitation (unit: mm/24 h, symbol Pt) is used to indicate precipitation factors in typhoons and rainstorms.
4. In additionally, we added three computable elements, namely precipitation duration (unit: d, symbol Pc), no precipitation duration (unit: d, symbol Sc), the number of days of high temperature (unit: d, symbol Th) according to the specific weather process affecting dengue.

By the above methods, we broke down a weather process into different weather indexes, which made the calculation of weather process influence on dengue fever become possible. We classified weather processes from 2005 to 2016 by weather indexes (Daily average temperature[T, °C], Daily average relative humidity[τ, %], Daily precipitation[Pt, mm], Continuous no precipitation days[Sc, d], Continuous precipitation days[Pc, d], Continuous high temperature days[Th, d]) in **Table 1**, and to further establish the prediction formula.

**Table 1.** Classification of meteorological indexes based on the features of weather process from 2005.01.01 to 2016.12.31.

| **Year** | **Daily average temperature ( T, ℃ )** | **Daily average relative humidity ( τ, % )** | **Daily precipitation ( Pt, mm )** | **Continuous no precipitation days ( Sc, d )** | **Continuous precipitation days ( Pc, d )** | **Continuous high temperature days ( Th, d )** |
| --- | --- | --- | --- | --- | --- | --- |
| **2005** | < 23 | < 30 | 0 | 1 | 0 | 1 |
| **2006** | 23-24 | 30 -35 | 0-1 | 2 | 1 | 2 |
| **2007** | 24-25 | 35 -40 | 1-5 | 3 | 2 | 3 |
| **2008** | 25-26 | 40 -45 | 5-10 | 4 | 3 | 4 |
| **2009** | 26-27 | 45 -50 | 10-20 | 5 | 4 | 5 |
| **2010** | 27-28 | 50 -55 | 20-30 | 6 | 5 | 6 |
| **2011** | 28-29 | 55 -60 | 30-40 | 7 | 7 | 7 |
| **2012** | 29-30 | 60 -65 | 40-50 | 8 | 8 |  |
| **2013** | 30-31 | 65 -70 | >50 | 9 | 9 |  |
| **2014** | 31-32 | 70 -75 |  | 13 | 10 |  |
| **2015** | 32-33 | 75 -80 |  | 15 | 11-20 |  |
| **2016** | > 33 | 80 -85 |  | 16 | >21 |  |

### 7. Statistical method

Part data were presented as mean±SD using SPSS 20.0 software (SPSS Inc., Chicago, IL, USA)

## Results

### 1 Overal temporal and spatial distribution characteristics of dengue cases

From January 1, 2005 to December 31, 2016, the total number of dengue cases in Guangzhou was 46,206 with 343 imported cases. It is worth noting that 45,870 dengue cases were reported from July to November, accounting for 99.28% of the total cases. From January to June, only 335 cases (0.73%) were reported, and February had the lowest incidence during a whole year **(Figure 1A)**.

**Figure 1.**
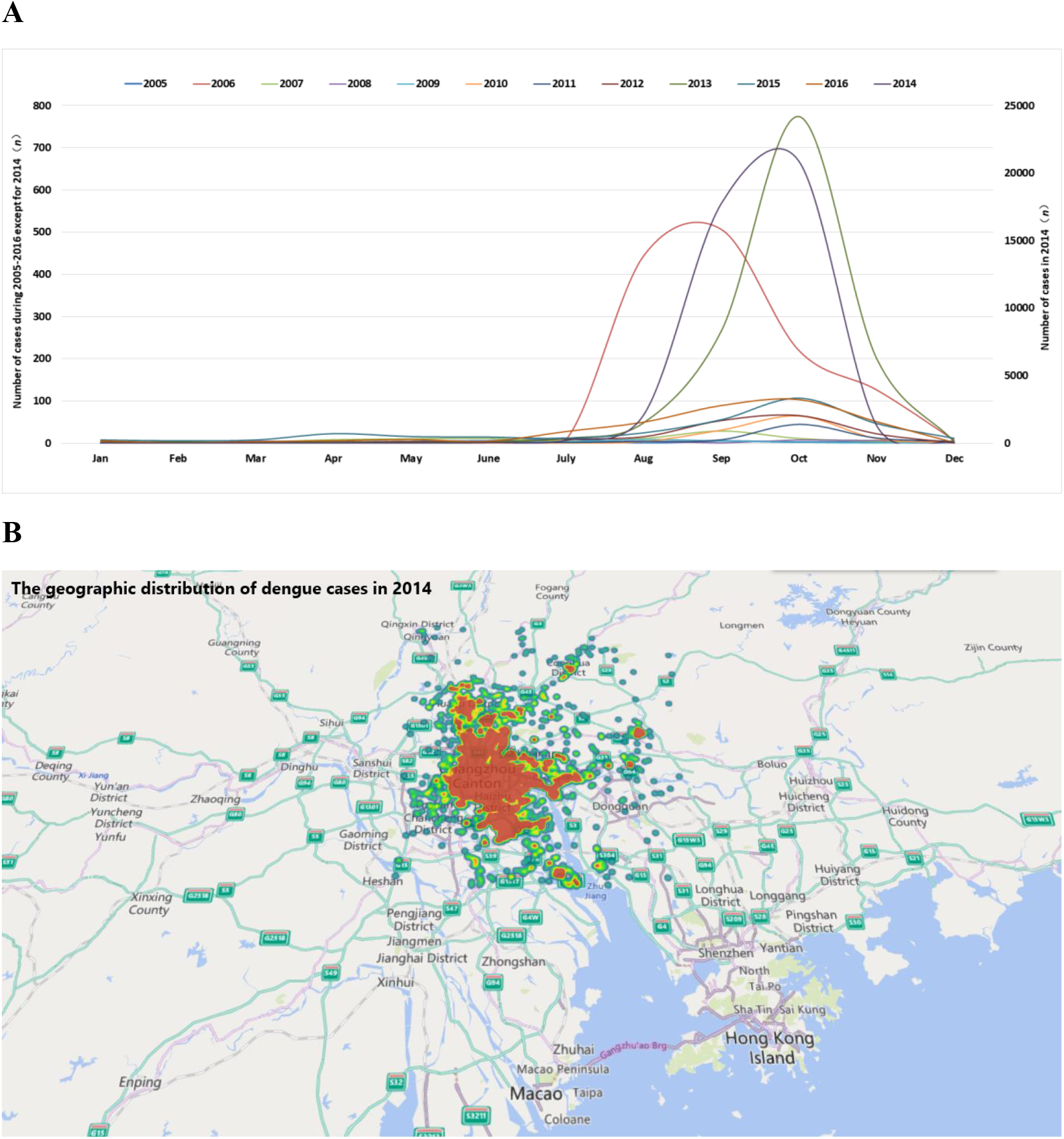
Temporal and spatial distribution characteristics of dengue cases from 2005 to 2016. A: Time distribution of dengue cases from 2005 to 2016. The right ordinate shows the case number of dengue for all years except 2014. B: Accumulatively spatial distribution of dengue cases from 2005 to 2016 in Guangzhou.

2014 had the most serious epidemic of the 12 years with 42,335 cases reported, accounting for 91.62% cases of the 12 years (**Figure 1B**); 2006 and 2013 had 1,319 cases and 1,311 cases, respectively, accounting for 2.85% and 2.84%, respectively. There were no more than 500 cases per year in the rest 9 years.

### 2 Overal temporal and spatial distribution characteristics of imported dengue cases

From January 1, 2005 to December 31, 2016, 343 cases were imported, accounting for 0.74% of the total number of cases (**Figure 2A**). Among them, 86 cases were imported in 2014, followed by 2015 (72 cases) and 2016 (56 cases). **Figure 2B** shows the ratio of imported cases of total cases in each year from 2005 to 2016. And in part years, the ratio of imported case was very high (2005, 24.24%; 2009, 57.14%; 2010, 23.17%, 2015, 22.29%, 2016, 15.73%).

**Figure 2.**
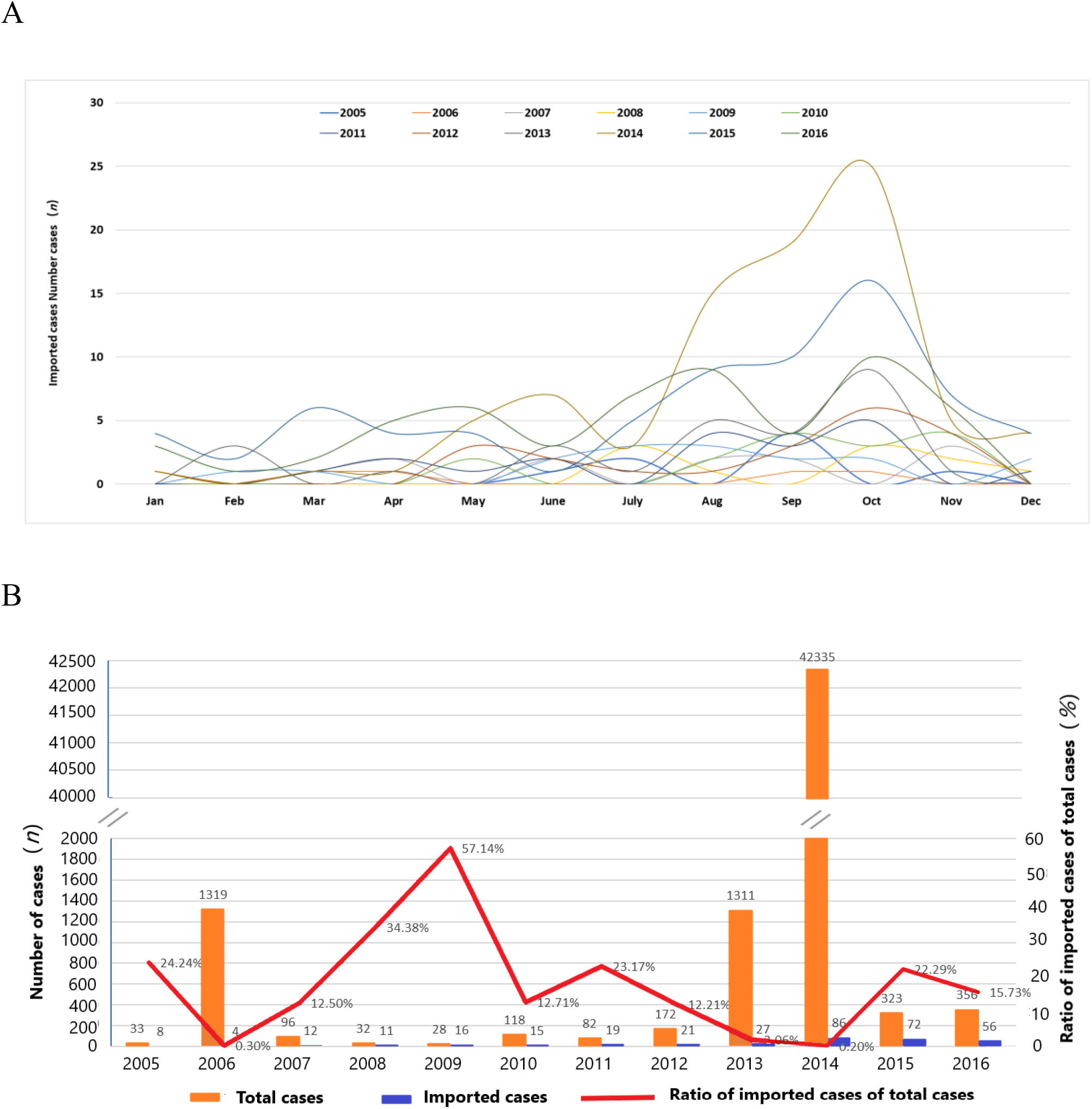
Temporal and spatial distribution characteristics of imported dengue cases from 2005 to 2016. A: Time distribution of dengue imported cases. B: The ratio of imported cases of total cases in each year.

From May to November is the main period for cases import, the number of cases imported reached 289, accounting for 83.29% of the total imported cases. The maximum number of imported cases was 80 in October, accounting for 23.05% of all the imported cases. The lowest import was in February, with a total of 7 imported cases, or 2.02% of the total imported cases. In the 12 years from 2005 to 2016, the first imported case occurred on January 1 at the earliest and June 30 at the latest.

### 3. The influence of imported cases on the epidemic of dengue

The correlation of imported cases on dengue epidemic in Guangzhou city was carried out. Under the confidence of α=0.01, the correlation coefficient reached 0.891, indicating that there was a positive correlation between the imported cases and the occurrence of dengue in Guangzhou.

By **Formula 1**, the distance of new cases from the imported case was evaluated. We set the imported case as the center, the horizontal axis was the distances from the case, and the vertical axis was the number of cases within different days after the onset time. The influence of imported cases on the around epidemic of dengue cases was evaluated in **Figure 3**. The results showed that the high-density area for case distribution is 5,000 meters to 6,500 meters on distance and within 5 days on time, with a potential dengue case incidence of 0.0181%. The area on the distance within 1,000 meters and on time longer than 15 days shows a low-density area of cases reported. It is generally believed that the range of the mosquito’s activity is mainly within 100 m of its birthplace, and the maximum is no more than 1 km(*12*), and the dengue incubation in human body is 3-10 days(*14*), so the area is with most influence of the imported case, but the result showed that the potential dengue cases incidence is only 0.0021% (**Figure 3**).

**Figure 3.**
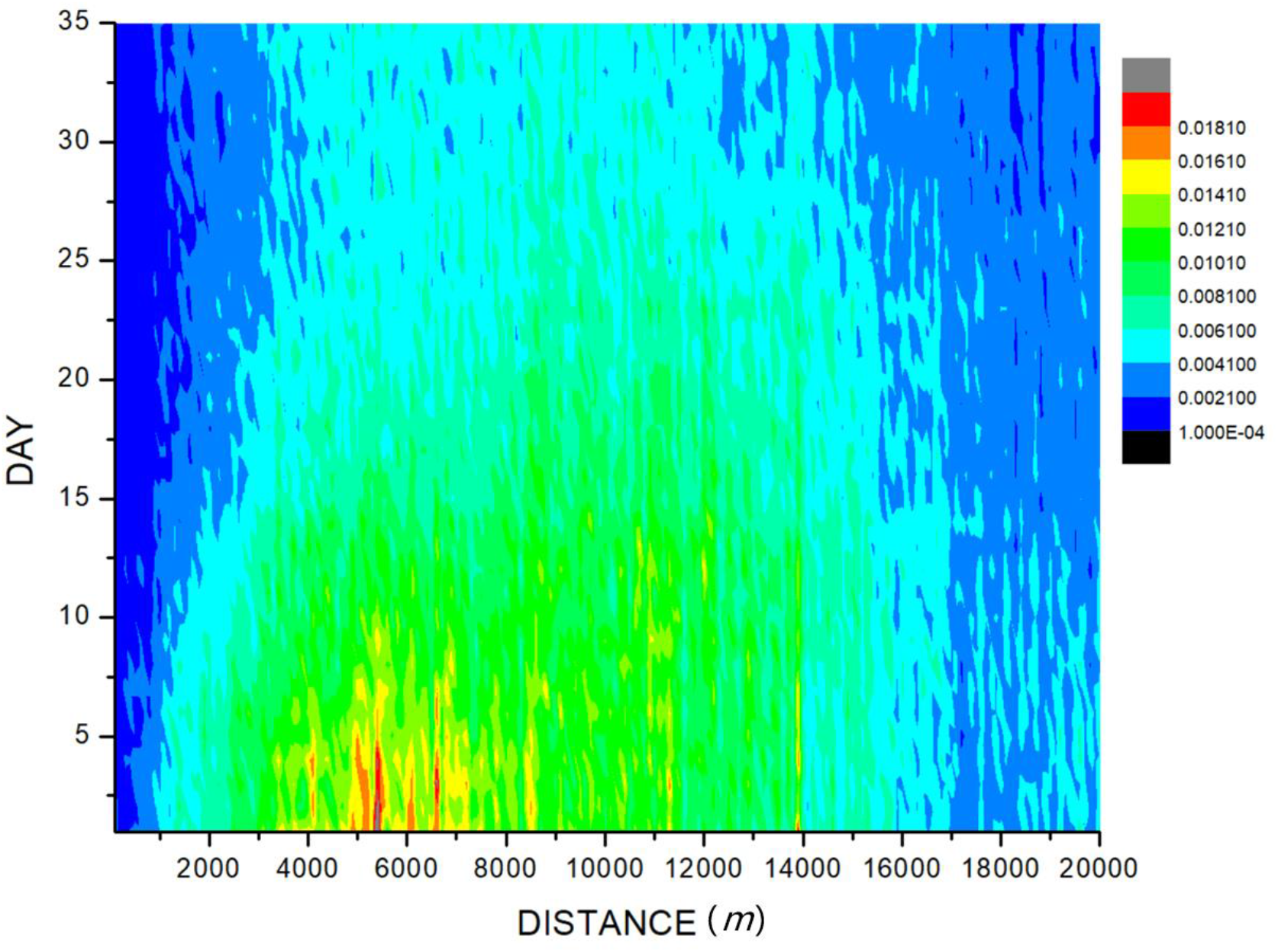
The influence of imported cases on the epidemic of surrounding dengue. The imported case was set as the original point, and the high-density area for case distribution is 5,000 meters to 6,500 meters from the imported case on distance and within 5 days on time of diagnosis of the imported case with a potential dengue incidence of 0.0181%. The area on the distance within 1,000 meters and on time longer than 15 days shows a low-density area of cases reported with the potential dengue incidence of only 0.0021%.

## 4. Establishment of prediction system of dengue

### 4.1 Prediction of the dengue season by a sliding accumulated temperature method

We calculated the average days between the date of the first case reported and the predicted date of T*n* ≥ 0, when *n* is 90 days (T_90_), 60 days (T_60_), 45 days (T_45_), 35 days (T_35_), 20 days (T_20_), 10 days (T_10_), and 6 days (T_6_), respectively, from 2005-2016 (**Table 2**).

**Table 2.** The average days between the date of the first case reported and the predicted date under various sliding accumulated temperature

| **T<sub>n</sub> ( °C )** | **The average days between the two dates (mean± SD)** | **T<sub>n</sub> ( °C )** | **The average days between the two dates (mean± SD)** |
| --- | --- | --- | --- |
| **T<sub>90</sub>≥0** | 13.55 ±28.44 | **T<sub>90</sub><0** | 55.25 ±31.93 |
| **T<sub>60</sub>≥0** | 4.64 ±28.60 | **T<sub>60</sub><0** | 29 ±22.88 |
| **T<sub>45</sub>≥0** | -1.55 ±27.38 | **T<sub>45</sub><0** | 19.92 ±22.86 |
| **T<sub>35</sub>≥0** | -9.55 ±24.80 | **T<sub>35</sub><0** | 16.42 ±22.44 |
| **T<sub>20</sub>≥0** | -18.82 ±26.05 | **T<sub>20</sub><0** | 6.75 ±23.92 |
| **T<sub>10</sub>≥0** | -22 ±25.14 | **T<sub>10</sub><0** | 1.91 ±23.79 |
| **T<sub>6</sub>≥0** | -23.09 ±23.83 | **T<sub>6</sub><0** | -0.67 ±23.72 |

We found that when the T_45_≥0, the average days between the two dates is -1.55 days, which is minimum with a standard deviation of 27.38 days, indicating that T45≥0 °C was an effective indicator of the beginning of the dengue season. We also calculated the average days between the date of the last case reported and the date with T_*n*_ <0, and found that when the sliding accumulated temperature is 6-day, the average date between the two dates is -0.67 days, which is the minimum with a standard deviation of 23.719, so we concluded that T_6_<0 °C had a valid indicative meaning for the ending of dengue fever season. **Formula 4** and **5** was derived as follows for T_45_ and T_6_ calculating:

#### Formula 4

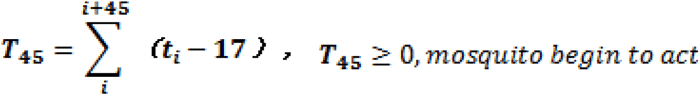

#### Formula 5

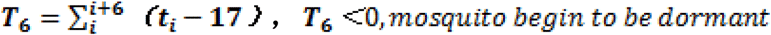

We chose T_45_ to set the predictive model, calculated the date of the first dengue case under T_45_ by **Formula 4** from 2005 to 2016, and verified the predicted date by the actual incidence date, which excluded the imported cases (**Table 3**). Calculating the accumulated cumulative temperature over 45 days, the date, which is mostly closed to the date of T_45_=0, and the number of cases accumulated in the 45 days before the special calculated day are compared in **Table 3**. The results showed that almost all dengue cases occurred after date with T_45_≥0 in Guangzhou, which means the T_45_ is an effective index for beginning of the dengue season.

**Table 3.**
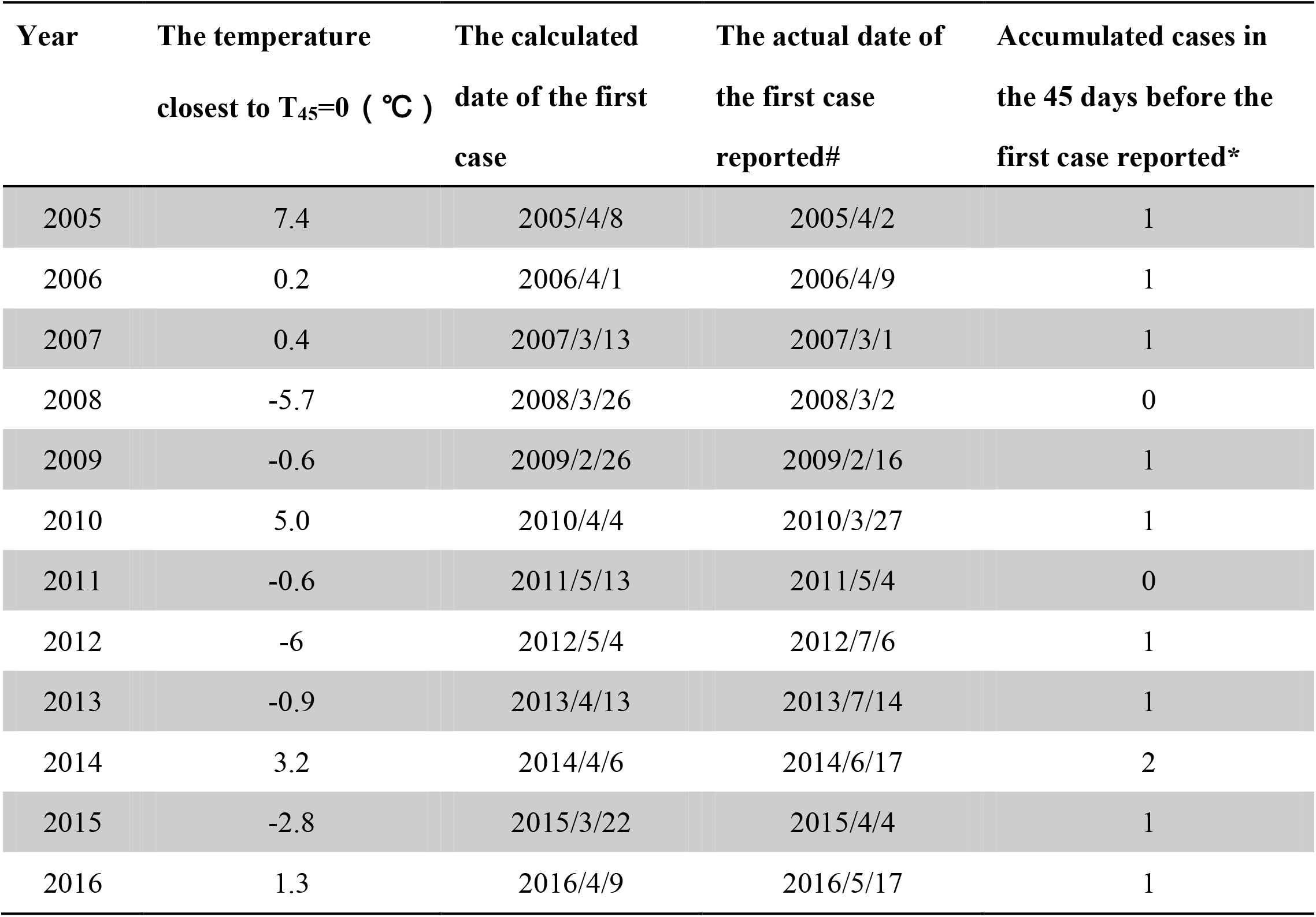
The calculated date of the first case by the sliding accumulated temperature method under T_45_ from 2005 to 2016

| **Year** | **The temperature**<br>**closest to T<sub>45</sub>=0 ( °C )** | **The calculated**<br>**date of the first**<br>**case** | **The actual date of**<br>**the first case**<br>**reported#** | **Accumulated cases in**<br>**the 45 days before the**<br>**first case reported\*** |
| --- | --- | --- | --- | --- |
| 2005 | 7.4 | 2005/4/8 | 2005/4/2 | 1 |
| 2006 | 0.2 | 2006/4/1 | 2006/4/9 | 1 |
| 2007 | 0.4 | 2007/3/13 | 2007/3/1 | 1 |
| 2008 | -5.7 | 2008/3/26 | 2008/3/2 | 0 |
| 2009 | -0.6 | 2009/2/26 | 2009/2/16 | 1 |
| 2010 | 5.0 | 2010/4/4 | 2010/3/27 | 1 |
| 2011 | -0.6 | 2011/5/13 | 2011/5/4 | 0 |
| 2012 | -6 | 2012/5/4 | 2012/7/6 | 1 |
| 2013 | -0.9 | 2013/4/13 | 2013/7/14 | 1 |
| 2014 | 3.2 | 2014/4/6 | 2014/6/17 | 2 |
| 2015 | -2.8 | 2015/3/22 | 2015/4/4 | 1 |
| 2016 | 1.3 | 2016/4/9 | 2016/5/17 | 1 |

We calculated the date of the last dengue case under T_6_ by **Formula 5** from 2005 to 2016, and verified the predicted date by the actual incidence date, which excluded the imported cases (**Table 4**). Calculating the accumulated cumulative temperature over 6 days, the date, which is mostly closed to the date of T_6_=0, and the number of cases accumulated after the special calculated day are compared in **Table 4**. The results showed that no more than one dengue cases occurred after date with T_6_<0, which means the T_6_ is an effective index for ending of the dengue season.

**Table 4.** The calculated date of the last case by the sliding accumulated temperature method under T_6_ from 2005 to 2016

| **Year** | **The accumulated temperature closest to $T_6=0$ ( °C )** | **The calculated date of the last case** | **The actual date of the last case reported#** | **Accumulated cases after the last case reported\*** |
| --- | --- | --- | --- | --- |
| 2005 | -2.5 | 2005/12/6 | 2005/11/9 | 0 |
| 2006 | -4.7 | 2006/12/14 | 2006/12/20 | 1 |
| 2007 | -0.2 | 2007/12/26 | 2007/12/19 | 0 |
| 2008 | -1.6 | 2008/11/30 | 2008/12/26 | 1 |
| 2009 | 3.3 | 2009/12/16 | 2009/12/26 | 1 |
| 2010 | 0.1 | 2010/1/12 | 2010/12/28 | 0 |
| 2011 | 0.6 | 2011/1/3 | 2011/12/5 | 0 |
| 2012 | 0.7 | 2012/12/29 | 2012/11/24 | 0 |
| 2013 | 1.8 | 2013/11/28 | 2013/12/12 | 1 |
| 2014 | 1.4 | 2014/12/5 | 2014/12/24 | 1 |
| 2015 | -2 | 2015/11/29 | 2015/11/29 | 0 |
| 2016 | -3.1 | 2016/11/26 | 2016/12/12 | 1 |

### 4.2 Establishment of prediction formula using meteorological parameters

By analysis of the occurrence of dengue cases under various weather process, we calculated the weights of each weather process and derived the formula of occurrence probability of dengue (Q) under a particular weather process (**Formula 6**):

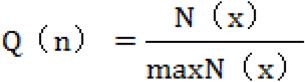

Where Q(n) is the probability constant corresponding to the particular weather process, x is the corresponding weather process, and N is the number of cases under the weather process with different corresponding level, so we established the **Formula 7** for dengue prediction under comprehensive consideration of weather processes:

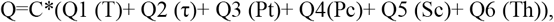

among which C is a constant about season factor, and Q1-Q6 is the probability constant calculated by the dengue case, and the parameter classification determines the probability constant by calculating the proportion of corresponding cases in the total cases.

C is related to the weather season, when Guangzhou is in the non-dengue season, C keeps small, when Guangzhou is in dengue season (July to Nov) or the weather meets the condition for dengue occurs and develops, C increases. **Formula 8** is used to calculate C

#### Formula 8

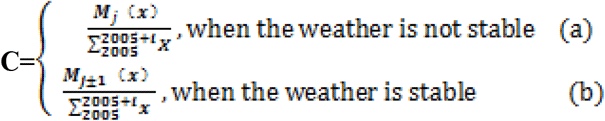

Where M is one of twelve months, *i* is the *i* year after 2005, and *j* is the arrange of month in twelve months, *x* is the case number about the M month of all years, *X* is all of the case number in 2005 to 2005+ *i* years. and when the weather is not stable, the C is calculated as formulate (a), and when the weather is stable, C will be the same as the C in previous or following month as formulate (b).

### 4.3 Forecast effects of dengue prediction formula

We got interpolations of various meteorological factors from in the third section “The relationship between dengue and meteorological factors”, and the interpolations of meteorological element data were substituted to the Probability Forecast (**Formula 7)** for high-precision prediction of any grid point of Guangzhou city.

We set Guangzhou as a grid point, and used average meteorological data of Guangzhou to plot the incidence curve of dengue of three years (2006, 2013, and 2014) by the Probabilistic Forecast and verify it by the actual incidence. We chose the three years with more than 1,000 reported dengue cases in order to better reflect the fitting degree between predicted curve and actual incidence curve. The curve of daily occurrence probability of dengue cases and the actual number of daily cases in 2006, 2013, and 2014 are shown in **Figure 4**, and the results showed that the predicted curve by the Probabilistic Forecast is in concordance with the actual case occurrence trend.

**Figure 4.**
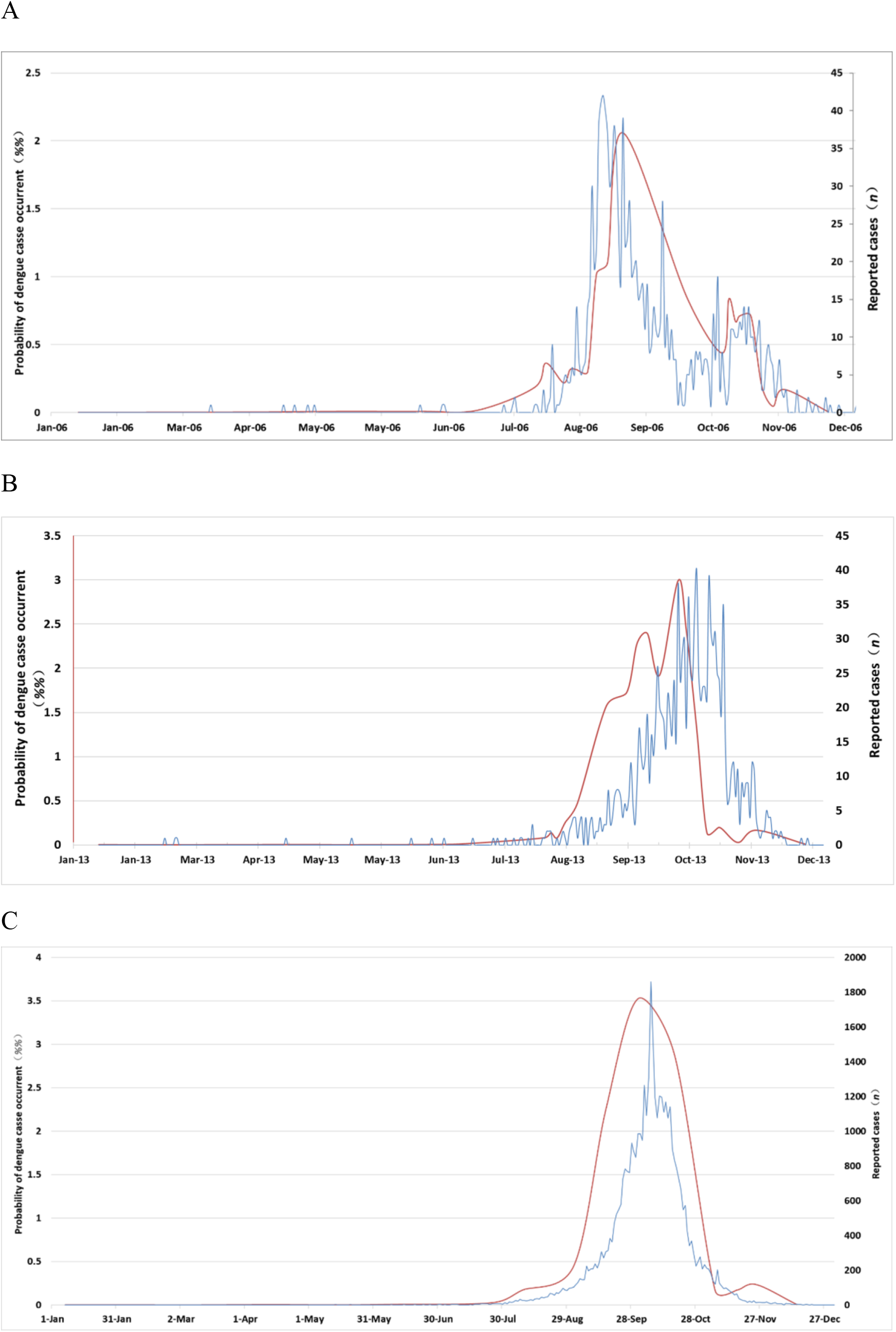
Comparison of prediction of dengue incidence curve (blue) and the actual curve of dengue cases (red). The red curve is the actual case curve of dengue fever, and uses the vertical axis on the right (unit: case number); The blue curve is the incidence prediction curve of dengue fever based on actual weather factors, uses the vertical axis on the left (unit: 10-4). A: The prediction of dengue incidence curve is significantly correlated with the actual curve in 2006, and the Correlation coefficient R is 0.751 with α of 0.01; B: The prediction of dengue incidence curve is significantly correlated with the actual curve in 2013, and the Correlation coefficient R is 0.896 with α of 0.01; C: The prediction of dengue incidence curve is significantly correlated with the actual curve in 2014, and the Correlation coefficient R is 0.873 with α of 0.01.

## Discussion

In this study, we established a Probabilistic Forecast model of dengue by cooperation of meteorology and dengue epidemiology in Guangzhou city. Using **Formula 4** derived from the temperature and humidity data, we accurately predicate the incidence of dengue in Guangzhou; That is, when T_45_≥0, the city enters the high-risk dengue season, and when T_6_ < 0, the city enters low-risk dengue season. Using **Formula 7**, we can predict the epidemic trend and degree of dengue. Combining **Formula 4, 5**, and **7**, we established a Probabilistic Forecast model of dengue epidemic, which can predict dengue incidence season with accurate beginning and ending date, and epidemiological status in the city of Guangzhou, where is the main area of dengue infection of China. The Probabilistic Forecast model can provide the government with accurate data for prevention and control of dengue, and help the making of effective strategies for full use of limited medical resources.

Since the relationship between climate information and dengue has been confirmed(*1, 15*), establishment of an early warning system for dengue epidemic is a research hotspot. Most of studies on early warning system used statistical methods to produce dengue predictions. Jain *et al*(*16*). used Generalized Additive Models (GAMs) to fit the relationships between the predictors and the clinical data of dengue from 2008 to 2012 in in Thailand, and evaluated the predictive ability of the fitted models according to RMSE and SRMSE, and concluded the model allows to make forecasts with a lead time of one month. Shi *et al*.(*17*) developed a set of statistical models using least absolute shrinkage and selection operator methods to forecast the weekly incidence of dengue notifications over a 3-month time. Lowe *et al*.(*18*) used seasonal climate and El Niño forecasts to predict the evolution of the dengue season in Machala, and the autoregressive model allows dengue in one calendar month to depend on dengue in the previous calendar month. Sang *et al*.(*11*) developed a time series Poisson multivariate regression model, and the results showed that imported cases in the previous month, monthly minimum temperature in the previous month and monthly accumulative precipitation with three-month lags could predict dengue outbreak ahead by one month.

Different from above studies, which used statistical methods to obtain probabilistic forecasting equation by study of climate factors and dengue cases through correlation, the following studies set up dengue prediction models by meteorological parameter. Lowe *et al*. performed several studies on the relationship between dengue and meteorological factors in Brazil, based on which they established an early warning system driven by real-time seasonal climate forecasts and the dengue cases reported to the Brazilian Ministry of Health in February 2014(*19-22*), however they used incomplete surveillance data to drive the model, the spatial resolution of forecasts were coarse, the information regarding the (re)introduction of different serotypes or vector control activities were lack. Lee *et al*(*23*) used a sensitivity analysis of the temperature-dependent parameters to explore the effects of climate change on dengue transmission dynamics.

The current study weighed the influence of meteorological indexes on dengue transmission during different weather processes, consequently determined the dengue parametric prediction equation with relative benefits and drawn against dengue vector hatching and large-scale breeding conditions. To our knowledge, **it is the first study** on parametric prediction model of dengue under comprehensive consideration of the influence of weather processes, including typhoons, heavy rain, high temperature, cold air mass, drought, and further determined the relationship between weather and dengue fever in a refined and quantitative way.

The weather process is continuous, so its impact on dengue is also continuous, therefore we aimed to determine the actual high correlation between dengue and weather process using continuous and quantitative meteorological factors, further to form a continuous prediction equation quantitatively of dengue fever. The advantages of our study mainly focus on the following three aspects: (1) by decomposing the weather processes into meteorological parameters, the influence of meteorology on dengue has been transformed from traditional qualitative analysis to quantitative analysis. Through quantitative analysis, a breakthrough has been made in the research on the influence of meteorology on dengue. Meteorological data can be used to quantitatively calculate the start and end dates of dengue fever each year, and can be used to quantitatively calculate whether the weather changes are suitable for the epidemic of dengue fever or not. (2) the forecast method was changed from single-factor static statistical forecast to multi-meteorological element continuous forecast. Traditional static statistical forecast can’t consider all the weather conditions, which means it can’t forecast the dengue under weather conditions that did not appear in history, so its application is limited. Our study is a multi-factor analysis, which predict the occurrence and development of dengue fever in the future under all weather conditions. It is a dynamic multi-factor continuous prediction equation, which can adjust the prediction results in real time with the change of actual weather conditions and the development of dengue fever.(3) The data accuracy is significantly improved, and the significance of prevention work guidance is: (a) the residential address of each case was precisely located; (b) By positioning, the meteorological data interpolation and every case location matched, so as to get the weather data and case highly matched, which ensured the basis and feasibility of meteorological elements and dengue epidemic research.

In this study, we developed a sliding accumulated temperature method to determine the beginning and ending date of high risk season of dengue. By the sliding accumulated temperature T_45_, we can calculate the accurate date of mosquito activity in Guangzhou area every year, after which local dengue fever is likely to start. The results would much benefit the local Prevention and Control Department to prevent and control dengue epidemic. This method is also firstly reported.

For the role of imported cases, we also got some interesting results. When we used the statistical way to analyze the correlation between imported cases and dengue epidemic in Guangdong Province, the results showed that the correlation coefficient reached 0.891 under the confidence of α=0.01, indicating that there was a positive correlation between the imported cases and the occurrence of dengue in Guangzhou. However, when we used the spatiotemporal method to analyze the influence of the imported case, we found that in the high-risk area of around the imported case (1000 m) within a period of 15 days, the potential dengue cases incidence is only 0.0021%, which means a very slight influence on incidence of surrounding dengue cases, and is inconsistent with the previous studies(*24*), in which, the authors concluded that this 2014 dengue outbreak in Guangzhou was initiated by an imported case from Southeast Asia based on the epidemiological results.

We found that the influence of imported cases on annual epidemic is closed to zero, even in the years with a high ratio of imported case (2005, 8/33[24.24%] ; 2009, 16/28[57.14%]; 2011, 19/82[23.17%], 2015, 72/323 [22.29%], 2016, 56 /356[15.73%]), all of these years had a total number of cases less than 500, so combined with the above information, we concluded that from 2005 to 2016, the sporadic single or a small number imported case of dengue may have a very slight effect on the area around which the patient lives and have not caused an epidemic of dengue around.

As we have confirmed that the imported cases had a positive correlation with the dengue epidemic in Guangzhou, the question comes, what is the potential role of imported case? We further analyzed the residence location and hospital admission distance and admission time of all valid imported cases through the spatiotemporal method, and found that within a radius of 0-20 km, the average distance from the imported case to the corresponding hospital was 5,519.9 meters, which was very close to the maximum impacting distance of the imported case (5.0-6.5 km).

As we have proved that sporadic single or a small number of imported case would not cause the epidemic, it provided us a bold guess that the hospitals receiving imported cases may be one of the key sites for dengue transmission, because hospital receiving the imported cases had the highest density of dengue patients, high flow of uninfected people with compared weak immunity, and open areas for mosquito.

In conclusion, we accurately calculated the beginning and ending day of dengue high risk season with the created sliding accumulated temperature method, and further successfully established a Probabilistic Forecast Model of dengue epidemic including number and density of incidence using meteorological factors with a good forecast effect, which was verified by the actual incidence data of dengue in Guangzhou from 2005-2016. The study provides evidence to public health department to generate strategy for prevention and control of dengue. The current Probabilistic Forecast Model is only for Guangzhou city because of the restriction of limited meteorological data, while the way to establish a valid Probabilistic Forecast Model of dengue by local meteorological data is worthy promoting among dengue endemic areas of the world.

Additionally, we confirmed that sporadic single or a small number of imported cases have a very slight influence on the dengue epidemic around, so high attention should be paid on the sites with concentrated patients, especially the hospitals receiving dengue patients, so the prevention and control policies should be regulated according to the new evidence.

## Data Availability

The data that support the findings of this study are available from the corresponding author (Jin Bu) upon reasonable request.

## 2. Author contributions

CJ provided overall guidance, MXF and DGH managed the project. BJ prepared the first draft and finished the manuscript on the basis of comments from other authors. All other authors provided data, developed models, reviewed results, provided guidance on methods, or reviewed the manuscript.

## Supplementary Materials

**Figure S1.**
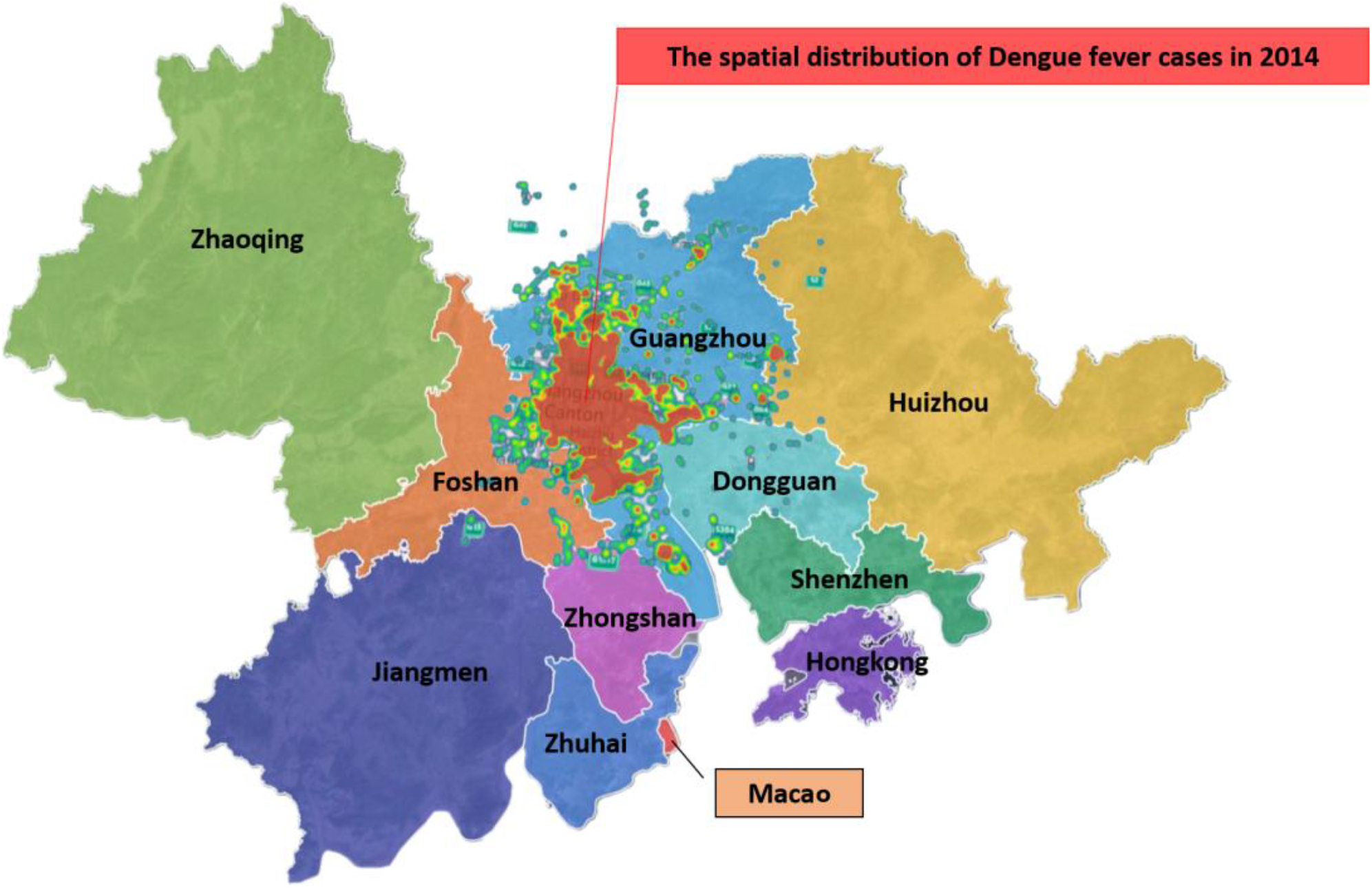
Spatial distribution of dengue case in 2014 in Guangzhou and its overlap with Guangdong-Hong Kong-Macao greater bay area.

## References

[1] Promprou S, Jaroensutasinee M, Jaroensutasinee K. Climatic factors affecting dengue haemorrhagic fever incidence in Southern Thailand. Dengue Bulletin, 2011, 29(1). doi:10.2004/wjst.v2i1.175.

[2] Bhatt S, Gething PW, Brady OJ, et al. The global distribution and burden of dengue. Nature 2013;496(7446):504–7. doi: 10.1038/nature12060.

[3] Liu K, Sun J, Liu X, et al. Spatiotemporal patterns and determinants of dengue at county level in China from 2005-2017. Int J Infect Dis 2018,77:96–104. doi: 10.1016/j.ijid.2018.09.003.

[4] Chen Y, Zhao Z, Li Z, et al. Spatiotemporal Transmission Patterns and Determinants of Dengue Fever: A Case Study of Guangzhou, China. Int J Environ Res Public Health 2019,16(14) doi: 10.3390/ijerph16142486.

[5] Alkhaldy I. Modelling the association of dengue fever cases with temperature and relative humidity in Jeddah, Saudi Arabia-A generalised linear model with break-point analysis. Acta Trop 2017;168:9–15. doi: 10.1016/j.actatropica.2016.12.034.

[6] Sanchez-Gonzalez G, Conde R, Noguez Moreno R,et al. Prediction of dengue outbreaks in Mexico based on entomological, meteorological and demographic data. PloS One 2018 Aug 6;13(8):e0196047. doi: 10.1371/journal.pone.0196047.

[7] Chien LC, Yu HL. Impact of meteorological factors on the spatiotemporal patterns of dengue fever incidence. Environ Int. 2014 Dec;73:46–56. doi: 10.1016/j.envint.2014.06.018.

[8] Couret J, Benedict MQ. A meta-analysis of the factors influencing development rate variation in Aedes aegypti (Diptera: Culicidae). BMC Ecol. 2014 Feb 5;14:3. doi: 10.1186/1472-6785-14-3.

[9] Mutheneni SR, Morse AP, Caminade C, et al. Dengue burden in India: recent trends and importance of climatic parameters. Emerg Microbes Infect. 2017 Aug 9;6(8):e70. doi: 10.1038/emi.2017.57.

[10] Hu W, Clements A, Williams G, et al. Spatial patterns and socioecological drivers of dengue fever transmission in Queensland, Australia. Environ Health Perspect 2012,120(2):260–266. doi: 10.1289/ehp.1003270.

[11] Dengue: Guidelines for Diagnosis, Treatment, Prevention and Control; Geneva: World Health Organization; 2009.

[12] Chan M, Johansson MA. The incubation periods of Dengue viruses. PLoS One. 2012;7(11):e50972. doi: 10.1371/journal.pone.0050972.

[13] Shang CS, Fang CT, Liu CM, et al. The role of imported cases and favorable meteorological conditions in the onset of dengue epidemics. PLoS Negl Trop Dis 2010,4(8):e775. doi: 10.1371/journal.pntd.0000775.

[14] Jain R, Sontisirikit S, Iamsirithaworn S, et al. Prediction of dengue outbreaks based on disease surveillance, meteorological and socio-economic data. BMC Infect Dis 2019,19(1):272. doi: 10.1186/s12879-019-3874-x.

[15] Shi Y, Liu X, Kok SY, et al. Three-Month Real-Time Dengue Forecast Models: An Early Warning System for Outbreak Alerts and Policy Decision Support in Singapore. Environ Health Perspect 2016,124(9):1369–1375. doi: 10.1289/ehp.1509981.

[16] Lowe R, Stewart-Ibarra AM, Petrova D, et al., Climate services for health: predicting the evolution of the 2016 dengue season in Machala, Ecuador. Lancet Planet Health. 2017 Jul;1(4):e142–e151. doi: 10.1016/S2542-5196(17)30064-5.

[17] Sang S, Gu S, Bi P, et al. Predicting unprecedented dengue outbreak using imported cases and climatic factors in Guangzhou, 2014. PLoS Negl Trop Dis 2015,9(5):e0003808. doi: 10.1371/journal.pntd.0003808.

[18] Lowe R, Barcellos C, Coelho CA, et al. Dengue outlook for the World Cup in Brazil: an early warning model framework driven by real-time seasonal climate forecasts. Lancet Infect Dis 2014,14(7):619–626. doi:10.1016/S1473-3099(14)70781-9.

[19] Lowe R, Bailey TC, Stephenson DB, et al. Spatio-temporal modelling of climate-sensitive disease risk: towards an early warning system for dengue in Brazil. Computers & Geosciences 2011,37, 371–381. doi: 10.1016/j.cageo.2010.01.008.

[20] Lowe R, Carvalho MS, Coelho CA, et al. Interpretation of probabilistic forecasts of epidemics. Lancet Infect Dis 2015,15(1):20. doi:10.1016/S1473-3099(14)71031-X.

[21] Lowe R, Coelho CA, Barcellos C, et al. Evaluating probabilistic dengue risk forecasts from a prototype early warning system for Brazil. Elife 2016,5 doi: 10.7554/eLife.11285.

[22] Lee H, Kim JE, Lee S, et al. Potential effects of climate change on dengue transmission dynamics in Korea. PLoS One, 2018;13(6):e0199205. doi: 10.1371/journal.pone.0199205.

[23] Peng HJ, Lai HB, Zhang QL, et al. A local outbreak of dengue caused by an imported case in Dongguan China. BMC Public Health 2012,12:83. doi: 10.1186/1471-2458-12-83.

[24] Zhong ZL, He GM. Life table of aedes albopictus at different temperatures. Journal of Sun Yat-Sen Medical University, 1988,35–39.(in Chinese)

